# Impact of systemic corticosteroids on hospitalized patients with COVID-19: January 2021 Meta-analysis of randomized controlled trials

**DOI:** 10.1101/2021.02.03.21251065

**Authors:** Robert Robinson, Vidhya Prakash, Raad Al Tamimi, Nour Albast, Basma Al-Bast

## Abstract

**Background:** The COVID-19 pandemic has stimulated worldwide investigation into a myriad of potential therapeutic agents, including corticosteroids. The first RCT reporting results on the impact of systemic corticosteroids on COVID-19 in a peer reviewed journal was the RECOVERY trial published in July, 2020. The RECOVERY trial showed a reduced risk of 28-day mortality in patients who received oral or intravenous dexamethasone for 10 days.

This study is a meta-analysis of peer reviewed RCTs aims to estimate the association of systemic corticosteroid therapy compared to the usual care or placebo on all-cause mortality in hospitalized patients with COVID-19. Software based tools to accelerate the analysis process.

**Methods:** Meta-analysis of peer reviewed RCTs comparing systemic corticosteroids to usual care or placebo.

**Results:** Five English language RCTs were identified, including data from 7645 hospitalized patients worldwide using systemic dexamethasone, hydrocortisone and methylprednisolone in COVID-19 positive patients. Three RCTs were discontinued when preliminary results from the RECOVERY trial became available.

Meta-analysis of all identified RCTs showed no difference in survival in patients who received systemic corticosteroid therapy compared to usual care or placebo (Odds ratio 0.82, 95% CI 0.64-1.05, p = 0.09). Subgroup analysis from the 1967 critically ill patients in the identified RCTs showed improved survival in patients who received systemic corticosteroid therapy (Odds ratio 0.67, 95% CI 0.51-0.87, p = 0.01).

**Conclusions:** This meta-analysis of randomized controlled trials published in peer-reviewed literature by January 1, 2021 showed reduced mortality in critically ill patients but not all hospitalized patients with COVID-19 who received systemic corticosteroids. The early termination of three of the included RCTs because of the preliminary results of the RECOVERY trial is likely to have dramatically influenced the results of this meta-analysis. Further research is needed to clarify the role of systemic corticosteroid therapy in the management of COVID-19.

## Introduction

The COVID-19 pandemic has stimulated worldwide investigation into a myriad of potential therapeutic agents. COVID-19 can cause highly elevated levels of inflammatory markers such as C-reactive protein, ferritin, GM-CSF, IL-1 and IL-6 ^1,2^. Corticosteroids can inhibit this inflammatory response at the cost of increasing the risk of nosocomial and opportunistic infections and worsening the course of COVID-19 ^3,4,5,6^. Because of these concerns, early guidelines for COVID-19 treatment stated that systemic corticosteroids are contraindicated or not recommended ^7,8^.

Observational and randomized controlled trials (RCTs) were immediately initiated to evaluate the impact of systemic corticosteroids on COVID-19. The first RCT reporting results in a peer reviewed journal, RECOVERY, was published in July, 2020 ^9^.

RECOVERY trial showed that 6mg of oral or intravenous dexamethasone for 10 days reduced the risk of 28-day mortality from 26% to 23% for all patients hospitalized due to COVID-19. Critically ill patients showed a greater mortality reduction from 41% to 29%. The positive results of the RECOVERY trial led to the suspension of most ongoing trials evaluating corticosteroids for the treatment of COVID-19.

A prospective meta-analysis conducted by the WHO Rapid Evidence Appraisal for COVID-19 Therapies (REACT) Working Group evaluating the role of systemic corticosteroids in the treatment of COVID-19 was published on October 6, 2020^10^. The REACT meta-analysis included data from the RECOVERY trial and partial data from 6 other RCTs showing a summary odds ratio of 0.66 (95% CI 0.53-0.82, p < 0.001). This indicates a reduced risk of death in patients with COVID-19 treated with systemic corticosteroids.

A more conventional meta-analysis by van Paassen et al. reviewed data from RCTs and observational studies^11^. It included data from 5 RCTs and 17 observational studies showing a summary odds ratio of 0.72 (95%CI 0.57–0.87, p = 0.002) indicating a reduced risk of death. The odds ratio with only RCTs was 0.84 (95%CI 0.69-0.99, p = 0.213), showing no clear impact on the risk of death in a pooled sample of 7645 patients.

This meta-analysis of peer reviewed RCTs aims to estimate the association of systemic corticosteroid therapy compared to the usual care or placebo on all-cause mortality in hospitalized patients with COVID-19. This study utilized software based tools to accelerate the analysis process.

## Methods

Data management and statistical analysis was conducted with R version 4.0.2. Meta-analysis calculations and plots were generated using the meta (version 4.15-1) and metafor (version 2.4-0) packages for R.

The protocol for this meta-analysis was registered and published in the PROSPERO database (CRD42020224991) on December 8, 2020^14^.

### Identification of trials

Trials were identified by searching MEDLINE using the search terms *COVID-19, corticosteroids*, and *steroids*. The results of these searches were exported and filtered via the RobotSearch tool by Vortext Systems to identify randomized controlled trials^12^. The filtered results were reviewed for inclusion criteria and evaluated for bias via the RobotReviewer tool that applies the Cochrane Risk of Bias (RoB) tool to the full text of articles^13^. ClinicalTrials.gov will also be searched to identify any other published trials.

Inclusion criteria for this study were: randomized controlled trials (RCTs) comparing systemic corticosteroids to other treatments or placebo in hospitalized adults with COVID-19 infections published by January 1, 2021 in an English language MEDLINE-indexed peer reviewed journal.

### Search Strings

((“2020/01/01” [Date -Publication] : “2021/01/01” [Date -Publication])) AND (steroid covid-19[Title])

((“2020/01/01” [Date -Publication] : “2021/01/01” [Date -Publication])) AND (corticosteroid covid-19[Title])

### Meta-analysis

A random-effects meta-analyses using the Paule-Mandel estimate of heterogeneity was used to determine odds ratios. The Hartung-Knapp adjustment was used to address uncertainty in the estimates of between-study variance in the random-effects meta-analysis.

Inconsistency in associations were evaluated using the I^2^ statistic and the Cochran Q statistic. Exact p values are reported.

Odds ratios and 95% confidence intervals were plotted for each trial to evaluate the association between corticosteroid therapy compared with usual care or placebo to the outcome measure.

Meta-analysis of the trials reporting data on critically ill patients included all trials where intensive care unit (ICU) admission was an eligibility criteria and the data on the patients who were on invasive mechanical ventilation (IMV) at randomization in all other trials.

Meta-analysis of trials reporting data on non-critically ill patients excluded all trials where ICU admission was an eligibility criteria and the data on the patients who were not on IMV at randomization in all other trials.

### Outcomes

The primary outcome evaluated for this meta-analysis was all cause mortality up to 30 days after randomization. Shorter timeframes were acceptable if 30 day survival data was not available.

## Results

The search strategy identified five English language RCTs with results published in peer reviewed literature (Figure 1). These 5 RCTs included data of 7645 hospitalized patients from Australia, Brazil, Canada, the European Union (EU), New Zealand, UK and the USA (Table 1). These studies used systemic dexamethasone, hydrocortisone and methylprednisolone in COVID-19 positive patients. These interventions were compared to placebo (2 RCTs) or usual care (3 RCTs). Three RCTs were discontinued when preliminary results from the RECOVERY trial became available.

**Table 1.** Included RCTs

| Table 1. Included RCTs |  |  |  |  |  |
| --- | --- | --- | --- | --- | --- |
|  | CAPE COVID | CoDex | METCOVID | RECOVERY | REMAP-CAP |
| ClinicalTrials.gov ID | NCT02517489 | NCT04327401 | NCT04343729 | NCT043881936 | NCT04343729 |
| Patients |  |  |  |  |  |
| Number | 149 | 299 | 393 | 6425 | 379 |
| Age (Mean) | 62 | 61 | 55 | 66 | 60 |
| Women | 30% | 37% | 35% | 36% | 29% |
| Confirmed COVID-19 | 97% | 96% | 81% | 89% | 72% |
| IMV at enrollment | 81% | 100% | 34% | 16% | 55% |
| Eligibility criteria |  |  |  |  |  |
| Known/Suspected COVID-19 | Yes | Yes | Yes | Yes | Yes |
| Acute respiratory failure | Yes | Yes | Yes | Yes | Yes |
| ICU Admission | Yes | Yes |  |  |  |
| IMV |  | Yes | Yes | Yes | Yes |
| Moderate/Severe ARDS |  | Yes |  |  |  |
| Intervention |  |  |  |  |  |
| Corticosteroid | Hydrocortisone | Dexamethasone | Methylprednisolone | Dexamethasone | Hydrocortisone |
| Dose | Continuous IV x8 or 14d<br>200mg/day x 4-7d<br>100mg/d x 4-7d<br>50mg/d 2-3d | 20mg IV daily x5d then 10mg IV daily x5d | 0.5 mg/kg IV 2x daily for 5 days | 6mg PO or IV daily x10d | 50mg IV every 6 h for 7 days |
| Control | Placebo | Usual Care | Placebo | Usual Care | Usual Care |
| Outcome | 21 day mortality | 28 day mortality | 28 day mortality | 28 day mortality | 21 day mortality |
| Setting | ICU | ICU | Hospital | Hospital | ICU |
| Location | France | Brazil | Brazil | UK | Australia, Canada, EU, New Zealand, UK, USA |
| Risk of bias |  |  |  |  |  |
| Random sequence generation | Yes | Yes | Yes | Yes | Yes |
| Allocation concealment | Yes | Yes | Yes | Yes | Yes |
| Blinding of participants and personnel | Yes | No | Yes | No | No |
| Blinding of outcome assessment | Yes | Yes | Yes | Yes | Yes |

**Figure 1.**
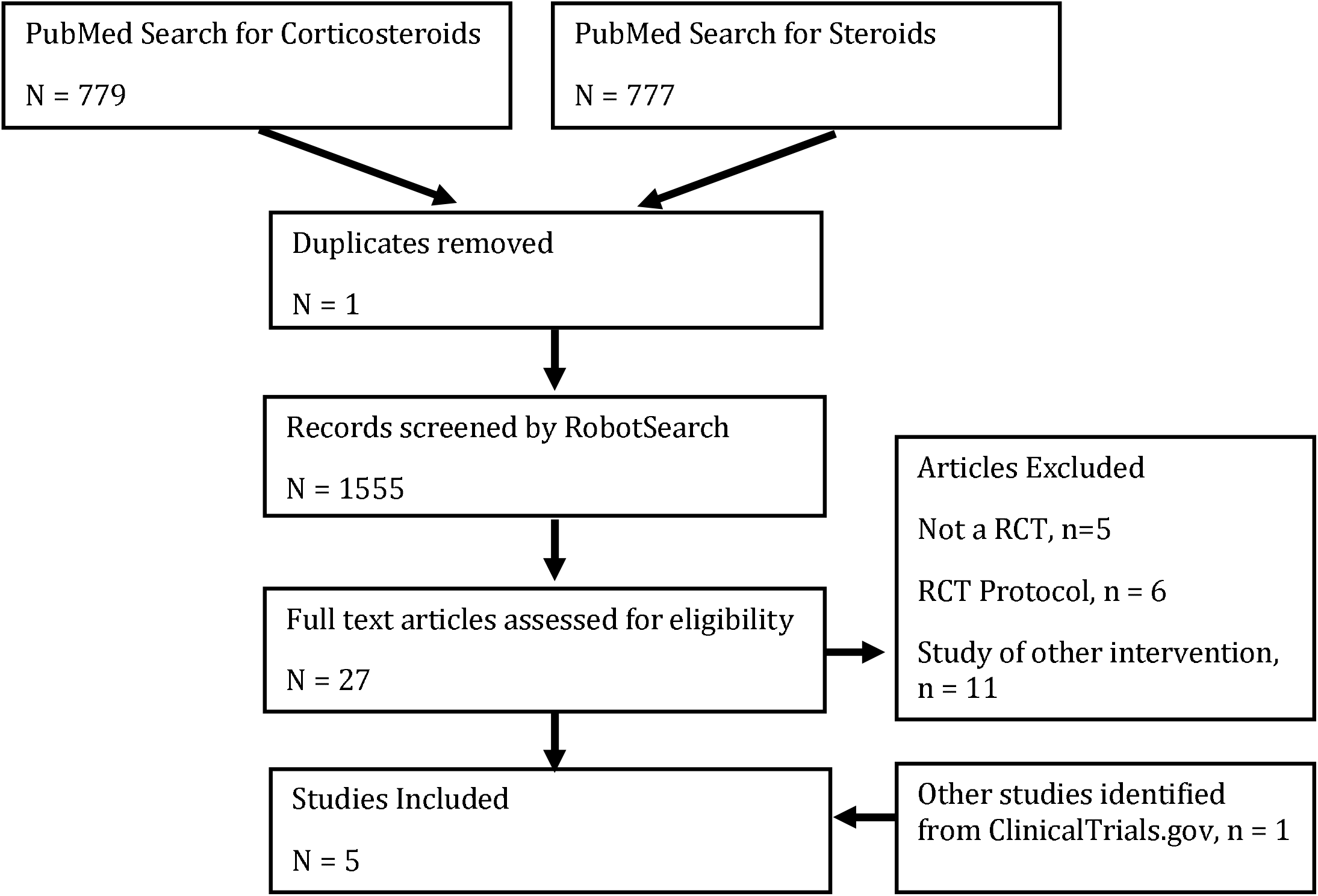
Article Selection Flowchart.

### CAPE COVID Trial

The CAPE COVID trial (NCT02517489) enrolled 149 critically ill patients with a high rate of invasive mechanical ventilation (IMV) at time of enrollment (81%) in a placebo-controlled trial to evaluate the impact of continuous intravenous hydrocortisone for 8-14 days on 21-day mortality. The average age in this study was 62 years and 30% of participants were women. COVID-19 infection was confirmed by laboratory testing in 97% of participants. The risk of bias in this placebo-controlled RCT was assessed to be low. This study showed no difference in mortality between the intervention and control groups (15% vs. 27%, p = 0.06). This trial was discontinued in June 2020 when preliminary data from the RECOVERY trial became available^15^.

### CODEX Trial

The CODEX trial (NCT04327401) enrolled 299 critically ill patients on IMV at time of enrollment in an RCT to evaluate the impact of intravenous dexamethasone for 10 days on 28-day mortality. The average age in this study was 61 years and 37% of participants were women. COVID-19 infection was confirmed by laboratory testing in 96% of participants.

This study has some risk of bias because treatment was not blinded. This study showed no difference in mortality between the intervention and control groups (56% vs.61%, p = 0.85). This trial was discontinued in June 2020 when preliminary data from the RECOVERY trial became available^16^.

### METCOVID Trial

The METCOVID trial (04343729) enrolled 393 hospitalized patients with a moderate rate of IMV at time of enrollment (34%) in a placebo-controlled trial to evaluate the impact of twice daily IV methylprednisolone for 5 days on 28-day mortality. The average age in this study was 55 years and 35% of participants were women. COVID-19 infection was confirmed by laboratory testing in 81% of participants. The risk of bias in this placebo-controlled RCT was assessed to be low. This study showed no difference in mortality between the intervention and control groups (37% vs.38%, p = 0.629)^17^. The METCOVID trial was not identified in the PUBMED search, but was found in searching ClinicalTrials.gov. METCOVID was PUBMED indexed at the time of our search.

### RECOVERY Trial

The RECOVERY trial (NCT043881936) enrolled 6425 hospitalized patients with a low rate of IMV at time of enrollment (16%) in an RCT to evaluate the impact of oral or intravenous dexamethasone for 10 days on 28-day mortality^9^. The average age in this study was 66 years and 36% of participants were women. COVID-19 infection was confirmed by laboratory testing in 89% of participants. This study has some risk of bias because treatment was not blinded. This study showed a statistically significant difference in mortality between the intervention and control groups (23% vs.26%, p < 0.001).

### REMAP-CAP Trial

The REMAP-CAP trial (NCT02735707) enrolled 379 hospitalized patients with a moderate rate of IMV at time of enrollment (55%) in an RCT to evaluate the impact of intravenous hydrocortisone for 7 days on 21-day mortality^18^. The average age in this study was 60 years and 29% of participants were women. COVID-19 infection was confirmed by laboratory testing in 72% of participants. This study has some risk of bias because treatment was not blinded. This study showed no difference in mortality between the intervention and control groups (28% vs. 33%, p not reported). This trial was discontinued in June 2020 when preliminary data from the RECOVERY trial became available.

### Association between systemic corticosteroids and mortality in all identified RCTs

There were 728 deaths in the 2803 patients randomized to receive systemic corticosteroids compared with 1330 deaths in the 4842 patients randomized to the control groups. Critically ill patients and patients who were not critically ill at the time of randomization were included in this meta-analysis. The random effects meta-analysis has a summary odds ratio of 0.82 (95% CI 0.64-1.05, p = 0.09, Figure 2). Heterogeneity assessment shows an I^2^ of 0% (p = 0.60).

**Figure 2.**
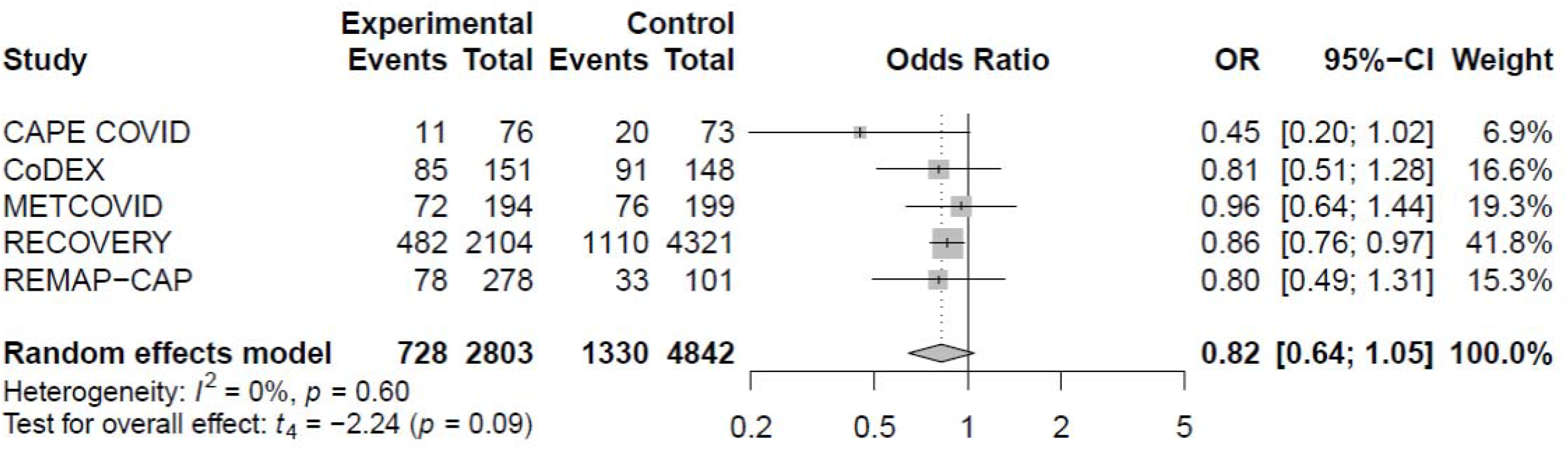
Association between Systemic Corticosteroids and All-Cause Mortality in All Identified RCTs.

### Association Between Systemic Corticosteroids and All-Cause Mortality In Critically Ill Patients In Identified RCTs

The CAPE-COVID, CODEX, and REMAP-CAP trials only included critically ill patients. The METCOVID and RECOVERY trial patients who were on IMV at randomization were included in this meta-analysis. In critically ill patients there were 322 deaths in the 895 patients randomized to receive systemic corticosteroids compared with 484 deaths in the 1072 patients randomized to the control groups. The random effects meta-analysis showed a summary odds ratio of 0.67 (95% CI 0.51-0.87, p = 0.01, Figure 3). Heterogeneity assessment shows an I^2^ of 0% (p = 0.58).

**Figure 3.**
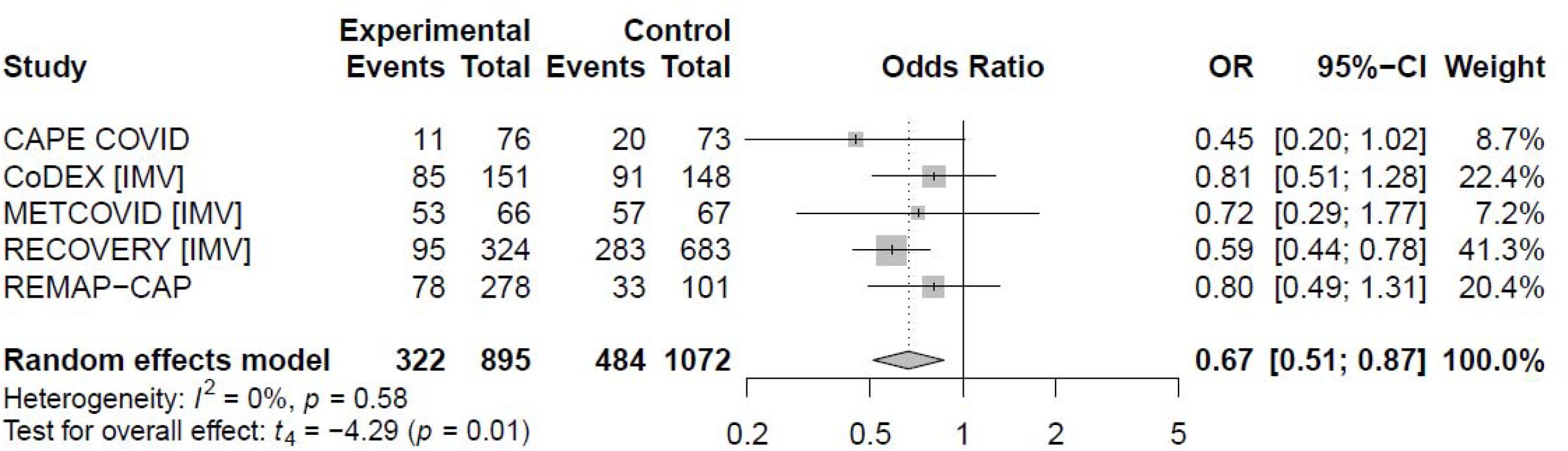
Association Between Systemic Corticosteroids and All-Cause Mortality In Critically Ill Patients In Identified RCTs.

### Association between Systemic Corticosteroids and All-Cause Mortality In Non-Critically Ill Patients In Identified RCTs

Only the METCOVID and RECOVERY trial included non-critically ill patients. In this population there was 406 deaths in the 1908 patients randomized to receive systemic corticosteroids compared with 846 deaths in the 3770 patients randomized to the control groups. The random effects meta-analysis showed a summary odds ratio is 0.95 (95% CI 0.75-1.19, p = 0.21, Figure 4). Heterogeneity assessment shows an I^2^ of 0% (p = 0.79).

**Figure 4.**
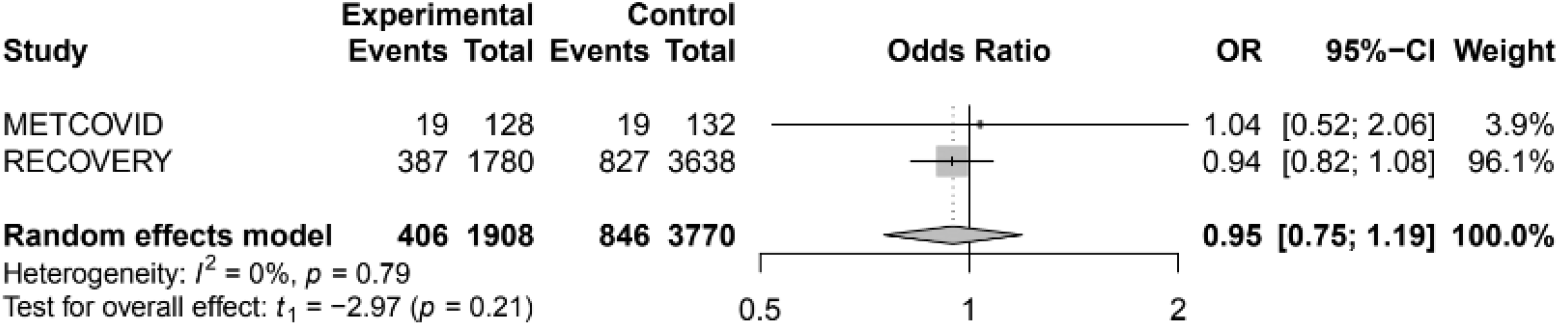
Association Between Systemic Corticosteroids and All-Cause Mortality In Non-Critically Ill Patients In Identified RCTs.

## Discussion

This meta-analysis of five RCTs included data of 7645 hospitalized patients from Australia, Brazil, Canada, the European Union (EU), New Zealand, UK and the USA. This study did not show a clear association between systemic corticosteroid treatment in COVID-19 patients and 28-day mortality in critically ill and non-critically ill patients.

Data from the 1967 critically ill patients in the identified RCTs showed improved survival in patients who received systemic corticosteroid therapy (Odds ratio 0.67, 95% CI 0.51-0.87, p = 0.01). These correspond well to the meta-analysis by Pasin et al. of 888 patients on IMV from the CODEX, METCOVID and RECOVERY trials (Odds ratio 0.85, 95%CI 0.72-1, p = 0.05)^19^.

Data from the non-critically ill patients in the METCOVID and RECOVERY trials showed no impact on mortality (Odds ratio 0.95, 95% CI 0.75-1.19, p = 0.21).

This study has several important limitations. The focus on English language publications in peer reviewed literature by January 1, 2021 can cause a significant bias by under-representing the full spectrum of research in this rapidly evolving field. Remarkably, these 5 trials were conducted and published within 9 months of the World Health Organization declaration of the COVID-19 pandemic on March 11, 2020. The results of additional RCTs evaluating the use of systemic corticosteroids in COVID-19 have been published on pre-print servers and it is reasonable to expect these and other studies to be published in peer-reviewed literature. Because of this rapid pace of research and the promising potential impact of systemic corticosteroids on outcomes in COVID-19 infection, this meta-analysis will be updated on a regular basis.

Treatment for COVID-19 has evolved considerably since the initiation of these studies. This can be clearly seen in the prevalence of other COVID-19 therapies outlined in table 2.

**Table 2.** Prevalence of other investigational COVID-19 therapies by RCT Blank cells indicate the use of this investigational COVID-19 therapy was not reported for the RCT.

| Table 2. Prevalence of other investigational COVID-19 therapies by RCT |  |  |  |  |  |
| --- | --- | --- | --- | --- | --- |
|  | CAPE COVID | CoDex | METCOVID | RECOVERY | REMAP-CAP |
| ClinicalTrials.gov ID | NCT02517489 | NCT04327401 | NCT04343729 | NCT043881936 | NCT04343729 |
| Azithromycin | 51 (34%) |  | 393 (100%) | 1581 (25%) |  |
| Eculizumab | 5 (3%) |  |  |  |  |
| Hydroxychloroquine | 70 (47%) |  |  | 39 (1%) |  |
| Lopinavir-ritonavir | 21 (14%) | 1 (<1%) |  | 6 (<1%) |  |
| Remdesivir | 5 (3%) |  |  | 5 (<1%) |  |
| Tocilizumab or Sarilumab | 3 (2%) |  |  | 171 (3%) |  |

The literature search and assessment methods used for this study were computer assisted, which differs from traditional meta-analysis techniques that are time and effort consuming. This strategy has the potential to introduce bias in the identification of RCTs. However, this methodology identified the same trials and did produce similar results (no statistically significant impact on survival) to a prospective meta-analysis conducted by the REACT Working Group and the van Paassen et al. meta-analysis ^10,11^. The failure to identify the METCOVID trial in the PUBMED search for this meta-analysis underscores the importance of including at least one other type of search in future meta-analysis projects.

The REACT prospective meta-analysis included data of 1703 patients, in comparison to this meta-analysis which included data of 7645 patients, due to the inclusion of complete enrollment and outcomes data from three RCTs that contributed interim results to the REACT meta-analysis and the results of an additional RCT. The results of this meta-analysis are similar to the results of the prospective meta-analysis (Odds ratio 0.82 (95% CI 0.64-1.05), p = 0.09 vs. Odds ratio 0.70, (95% CI 0.48-1.01), p = 0.053). Results of the DEXA-COVID-19 and Steroids-SARI studies included in the REACT meta-analysis were not published in the MEDLINE-indexed peer-reviewed literature by January 1, 2021.

The traditional search strategies in the van Paassen et al. meta-analysis identified the same studies identified in this meta-analysis. Results were nearly identical to the results in this study (Odds ratio 0.82 (95% CI 0.64-1.05), p = 0.09 vs. Odds ratio 0.84 (95% CI 0.69 – 0.99), p = 0.213). The slight differences between the results of these two meta-analyses is most likely due to differences in the software used for the analysis (R with the *meta* and *metafor* packages vs. Stata).

Systemic corticosteroid therapy showed a statistically significant reduction in mortality for critically ill patients (Odds ratio 0.67, p = 0.01) but not for all hospitalized patients (Odds ratio 0.82, p = 0.09). The termination of three of the five included RCTs after the preliminary results of the RECOVERY study were published is a significant confounding factor with these results. This may have led to an underestimation of the impact of systemic corticosteroids on mortality in hospitalized patients with COVID-19.

## Conclusions

This meta-analysis of randomized controlled trials published in peer-reviewed literature by January 1, 2021 showed reduced mortality in critically ill patients but not all hospitalized patients with COVID-19 who received systemic corticosteroids. The early termination of three of the included RCTs because of the preliminary results of the RECOVERY trial is likely to have dramatically influenced the results of this meta-analysis. Further research is needed to clarify the role of systemic corticosteroid therapy in the management of COVID-19.

## Data Availability

Data in used for the analysis in the manuscript is available on request.

## References

1. Moore JB, June CH. Cytokine release syndrome in severe COVID-19. Science 2020;368:473–474.

2. De Luca, Giacomo, et al. “GM-CSF blockade with mavrilimumab in severe COVID-19 pneumonia and systemic hyperinflammation: a single-centre, prospective cohort study.” The Lancet Rheumatology 2.8 (2020): e465–e473.

3. Rhen T, Cidlowski JA. Antiinflammatory action of glucocorticoids—new mechanisms for old drugs. N Engl J Med. 2005;353(16):1711–1723. doi:10.1056/NEJMra050541

4. Russell CD, Millar JE, Baillie JK. Clinical evidence does not support corticosteroid treatment for 2019-nCoV lung injury. Lancet. 2020;395(10223):473–475.

5. Ye Z, Wang Y, Colunga-Lozano LE, et al. Efficacy and safety of corticosteroids in COVID-19 based on evidence for COVID-19, other coronavirus infections, influenza, community-acquired pneumonia and acute respiratory distress syndrome: a systematic review and meta-analysis. CMAJ. 2020;192(27):E756–E767. doi:10.1503/cmaj.200645

6. Li H, Chen C, Hu F, et al. Impact of corticosteroid therapy on outcomes of persons with SARS-CoV-2, SARS-CoV, or MERS-CoV infection: a systematic review and meta-analysis. Leukemia. 2020;34(6):1503–1511. doi:10.1038/s41375-020-0848-3

7. Alhazzani W, Møller MH, Arabi YM, et al. Surviving Sepsis Campaign: guidelines on the management of critically ill adults with coronavirus disease 2019 (COVID-19). Crit Care Med. 2020;48(6):e440–e469. doi:10.1097/CCM.0000000000004363

8. Shang L, Zhao J, Hu Y, Du R, Cao B. On the use of corticosteroids for 2019-nCoV pneumonia. Lancet 2020;395:683–684.

9. Horby P, Lim WS, Emberson JR, et al; RECOVERY Collaborative Group. Dexamethasone in hospitalized patients with Covid-19—preliminary report. N Engl J Med. Published online July 17, 2020. doi:10.1056/NEJMoa2021436

10. The WHO Rapid Evidence Appraisal for COVID-19 Therapies (REACT) Working Group. Association Between Administration of Systemic Corticosteroids and Mortality Among Critically Ill Patients With COVID-19: A Meta-analysis. JAMA. 2020;324(13):1330–1341. doi:10.1001/jama.2020.17023

11. van Paassen, J., Vos, J.S., Hoekstra, E.M. et al. Corticosteroid use in COVID-19 patients: a systematic review and meta-analysis on clinical outcomes. Crit Care 24, 696 (2020). https://doi.org/10.1186/s13054-020-03400-9

12. Marshall, IJ, Noel-Storr, A, Kuiper, J, Thomas, J, Wallace, BC. Machine learning for identifying Randomized Controlled Trials: An evaluation and practitioner’s guide. Res Syn Meth. 2018; 9: 602– 614. https://doi.org/10.1002/jrsm.1287

13. Marshall IJ, Kuiper J, Wallace BC, RobotReviewer: evaluation of a system for automatically assessing bias in clinical trials, Journal of the American Medical Informatics Association, Volume 23, Issue 1, January 2016, Pages 193–201, https://doi.org/10.1093/jamia/ocv044

14. Robinson R and Prakash V. Impact of systemic corticosteroids on mortality in COVID-19. PROSPERO 2020 CRD42020224991 Available from: https://www.crd.york.ac.uk/prospero/display_record.php?ID=CRD42020224991

15. Dequin PF, Heming N, Meziani F, et al. Effect of hydrocortisone on 21-day mortality or respiratory support among critically ill patients with COVID-19: a randomized clinical trial. JAMA. Published online September 2, 2020. doi:10.1001/jama.2020.16761

16. Tomazini BM, Maia IS, Cavalcanti AB, et al. Effect of dexamethasone on days alive and ventilator-free in patients with moderate or severe acute respiratory distress syndrome and COVID-19: the CoDEX randomized clinical trial. JAMA. Published online September 2, 2020. doi:10.1001/jama.2020.17021

17. Jeronimo CMP, Farias MEL, Val FFA, et al. Methylprednisolone as adjunctive therapy for patients hospitalized with COVID-19 (Metcovid). Clin Infect Dis. Published online August 12, 2020. doi:10.1093/cid/ciaa1177

18. The Writing Committee for the REMAP-CAP Investigators. Effect of hydrocortisone on mortality and organ support in patients with severe COVID-19: the REMAP-CAP COVID-19 Corticosteroid Domain randomized clinical trial. JAMA. Published online September 2, 2020. doi:10.1001/jama.2020.17022

19. Pasin L, Navalesi P, Zangrillo A, Kuzovlev A, Likhvantsev V, Hajjar LA, Fresilli S, Lacerda MVG, Landoni G. Corticosteroids for Patients With Coronavirus Disease 2019 (COVID-19) With Different Disease Severity: A Meta-Analysis of Randomized Clinical Trials. J Cardiothorac Vasc Anesth. 2021 Feb;35(2):578–584. doi:10.1053/j.jvca.2020.11.057. Epub 2020 Nov 28. PMID: 33298370; PMCID: PMC7698829.

